# Identifying markers of health-seeking behaviour and healthcare access in UK electronic health records

**DOI:** 10.1101/2023.11.08.23298256

**Authors:** S. Graham, J.L. Walker, N. Andrews, D. Nitsch, P.K.E. Parker, H. I McDonald

## Abstract

**Objective:** To assess the feasibility of identifying markers of health-seeking behaviour and healthcare access in UK electronic health records (EHR), for identifying populations at risk of poor health outcomes, and adjusting for confounding in epidemiological studies.

**Design:** Cross sectional observational study using the Clinical Practice Research Datalink (CPRD) Aurum pre-linked to Hospital Episode Statistics.

**Setting:** Individual-level routine clinical data from 13 million patients across general practices (GPs) and secondary data in England.

**Participants:** Individuals aged ≥66 years on 01/09/2019.

**Main outcome measures:** We used the Theory of Planned Behaviour (TPB) model and the literature to iteratively develop criteria for markers selection. Based on this we selected 15 markers: those that represented uptake of public health interventions, markers of active healthcare access/use and markers of lack of access/underuse. We calculated the prevalence of each marker using relevant lookback periods prior to index date (01/09/2019) and compared to national estimates. We assessed the correlation coefficients (phi) between markers with inferred hierarchical clustering.

**Results:** We included 1,991,284 individuals (mean age: 75.9 and 54.0% females). The prevalence of markers ranged from <0.1% (low-value prescriptions) to 92.6% (GP visits), and most were in line with national estimates; e.g., 73.3% for influenza vaccination in the 2018/2019 season, compared to 72.4% in national estimates. Screening markers e.g., abdominal aortic aneurysm screening were under-recorded even in age-eligible groups (54.3% in 65–69 year-olds vs 76.1% in national estimates in men). Overall, marker correlations were low (<0.5) and clustered into groups according to underlying determinants from the TPB model.

**Conclusion:** Overall, markers of health-seeking behaviour and healthcare access can be identified in UK EHRs. The generally low correlations between different markers of health-seeking behaviour and healthcare access suggest a range of variables are needed to capture different determinants of healthcare use.

**Strengths and limitations of this study:**

- This is the first known study in the UK that has identified proxies or markers of health-seeking behaviour or healthcare access.
- We utilised linked electronic health records from primary and secondary care so that a range of different health utilisation markers could be identified.
- We identified a large population of over 2 million individuals.
- For some of the markers (e.g., bone density scans), health need could not be entirely separated from health behaviour and access.
- Marker prevalences showed different patterns by age, and these findings might not be generalisable to younger age groups (<65 years).

## Background

Health-seeking behaviour can be defined as “any activity undertaken by a person believing [themselves] to be healthy, for the purpose of preventing disease or detecting it in an asymptomatic stage”^1^. Healthcare access can be defined as “the ability to obtain healthcare services such as prevention, diagnosis, treatment, and management of diseases, illness, disorders, and other health-impacting conditions”^2^. Healthcare professionals or researchers might be interested in identifying patients with a lack of health-seeking behaviour or healthcare access, since these individuals are likely to suffer from worse clinical outcomes. Health-seeking behaviour and healthcare access may also be a key confounder in observational studies, and failure to account for this may undermine the validity of results. This type of confounding is thought to have contributed to overestimates of the protective effect of influenza vaccinations against all-cause mortality in observational cohort studies^3^. Information on health-seeking behaviour and healthcare access can be collected prospectively through surveys or interviews; for example, in the English Longitudinal Study of Ageing study^4^. Typically, in routinely-recorded data such as electronic health records (EHRs) it is difficult to identify health-seeking behaviour and healthcare access since they are not directly recorded. Suitable markers would need to represent interactions with the healthcare system (i.e., healthcare utilisation), preferably with limited dependence on underlying health need. Behavioural scientists have a variety of models for explaining the determinants for healthcare utilisation. For example, the updated Theory of Planned Behaviour (TPB) model^5^ describes the psychological, physical, contextual, and sociodemographic determinants for healthcare utilisation. Psychological determinants include influences on the micro and macro level such as societal attitudes, but also personal prior experiences. Physical determinants are on the micro-level and include lifestyle factors such as drug consumption, body mass index and physical activity. Context determinants are on the macro-level and include potential external barriers such as recommendations from healthcare professionals or geopolitical influences. Sociodemographic determinants are on the micro-level and include individual characteristics such as sex, age and living arrangements. These models demonstrate that there are a range of different determinants and therefore many different markers are likely required to capture all the underlying influences.

Three recent studies in the United States (US)^6-8^ introduced adjusting for markers of health-seeking behaviour and healthcare access in observational research. However, it is not known to what extent suitable markers can be identified in UK EHR. This study aimed to identify markers of health-seeking behaviour and healthcare access in UK EHRs, compare their prevalence to available national estimates, and explore correlations between different markers. This study will focus on individuals aged over 65 years as health-seeking behaviour and healthcare access varies by age^9^ and because they have high morbidity and mortality^10^.

## Methods

### Data sources and population

We used the Clinical Practice Research Datalink (CPRD) Aurum pre-linked to Hospital Episode Statistics (HES) admitted patient care (APC). CPRD Aurum holds anonymised longitudinal primary care patient records collected from the EMIS® Health patient record system. At the time of data extraction (May 2022 release) this data included 1,491 currently contributing general practices for 13,300,067 currently contributing patients (19.83% of the UK population). 99% of the practices are in England and <1% are in Northern Ireland^11^. CPRD Aurum uses a combination of SNOMED, Read codes (Clinical Terms Version 3) and local EMIS codes that are each individually mapped to a unique “medcode”. Prescriptions are recorded using the NHS dictionary of medicines and devices, each are mapped to a unique “prodcode”. HES APC is a secondary care commissioning dataset that covers all NHS secondary care in England^12^. HES uses International Classification of Diseases 10th Revision (ICD-10) codes^13^ to record diagnoses and Classification of Interventions and Procedures (OPCS) codes^14^ to record procedures. Our study population included individuals in England aged 66 years or older on the 1 September 2019. We only included individuals with a GP practice registration start date before 1 September 2018 to allow for a minimum one-year pre-index period for marker identification.

### Marker selection

We used the Theory of Planned Behaviour model to define our aim of identifying healthcare utilisation driven by determinants other than physical and mental health. We developed candidate markers and formal criteria for marker selection, incorporating input from two clinical epidemiologists on UK clinical practice and data recording (DN and HIM). Candidate markers from the aforementioned US studies^6-8^ were tested against these criteria to iteratively make improvements to the criteria and identify additional potential markers. For all the markers identified from previous literature see **Supplementary Table 1**. The final criteria that were developed can be found in **Table 1** below. We selected fifteen markers that included abdominal aortic aneurysm (AAA) screening; breast cancer screening; bowel cancer screening; cervical cancer screening; influenza vaccination; pneumococcal vaccination; NHS health checks; prostate specific antigen (PSA) testing; bone density scans; low-value procedures; glucosamine use (low-value prescription); GP practice visits; did not attend (DNA) primary care visit; hospital visit for ambulatory care sensitive (ACS) condition; and blood pressure measurements. In general the criteria were a good fit for the markers, but there was some tolerance for minor deviations, particularly for accepting some influence of underlying health conditions (**Supplementary Table 2)**.

**Table 1.** Criteria used to assess inclusion of markers of health-seeking behaviour and healthcare access.

| # | Criteria | Explanation | Example of a marker that does not meet criteria |
| --- | --- | --- | --- |
| 1 | Should be currently or recently available in national clinical practice to all individuals (overall or by sex) at cohort entry. | Ensures that the denominator population (by sex) is eligible for each of the markers. | Shingles vaccination is currently recommended in the UK to all individuals turning 65 years (among others).<sup>1</sup> However, it was not historically available for all age-cohorts in this study due to the evolving age-based eligibility criteria since vaccine introduction in 2013. As a result, only selected age-cohorts would have had a period of age-based eligibility for shingles vaccination at the study index date, and this would not have been a universal marker for the study population. |
| 2 | Should be routinely recorded in the available data sources. | Ensures routine ascertainment of markers which is not dependent on other factors such as abnormal test results. | Vision and hearing tests are available through the NHS in the UK; however, most people get these tests at a private optician. Although opticians routinely send results to GPs, these may be uploaded as a PDF rather than coded in the patient's health record, particularly if no abnormality is found. |
| 3 | Should not be primarily dependent on underlying health needs. | Ensures that the determinants of healthcare utilisation are not primarily driven by underlying health conditions. | Adherence to medication could represent health-seeking behaviour and healthcare access; however, medication use is dependent on a diagnosed condition or health need. |

Note: Shingles vaccine was first made available to immunocompetent individuals aged 70 or 79 in 2013 in the UK, with a phased catch-up programme for individuals aged 70–79 years. In 2021, the programme introduced recombinant vaccination which can be given to people with immunosuppression. At the time of the study index date, shingles vaccine was available to all individuals aged 70–79 years. Shingles vaccination is currently (1 September 2023) recommended in the UK to all individuals turning 65 years, currently aged 70–79, or aged 50 and over with immunosuppression.

Some markers represented active health-seeking behaviour and healthcare access, such as uptake of recommended vaccinations. Other markers represented lack of health-seeking behaviour and healthcare access – such as DNA for primary care visits, and hospital visits for ACS conditions. ACS conditions are conditions for which effective community care can help prevent the need for hospital admission^15^. If an individual has a visit to hospital for an ACS condition, then we can presume that they had a lack of healthcare access or health-seeking behaviour as they were unable to or did not access care when their symptoms were less severe. Low-value procedures and low value prescriptions are those that the National Institutes of Health and Care Excellence recommended to no longer provide in UK clinical practice since they were deemed to have little or no benefit, whilst still incurring an avoidable cost^16,17^. We considered both to be indicators of active health-seeking behaviour or healthcare access from a patient perspective, as patients were receiving (non-recommended) care for their perceived needs.

### Marker operational definitions

The operational definition of each marker includes code lists and lookback periods to apply in the current study datasets (**Table 2**).

**Table 2.**
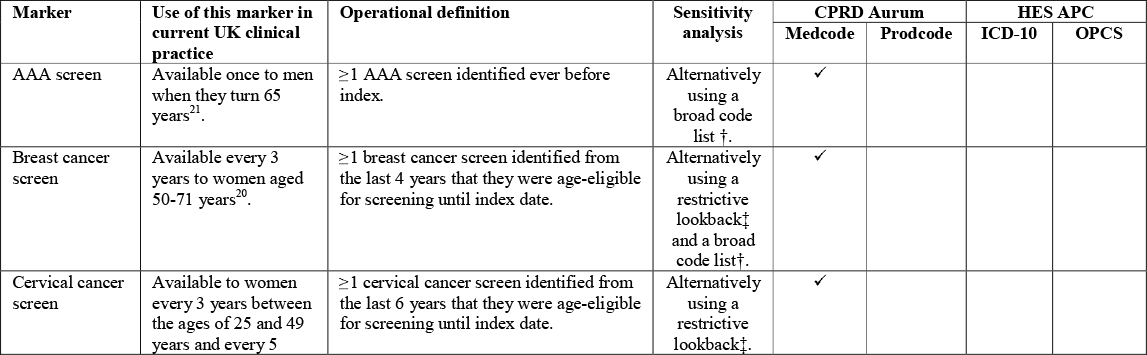
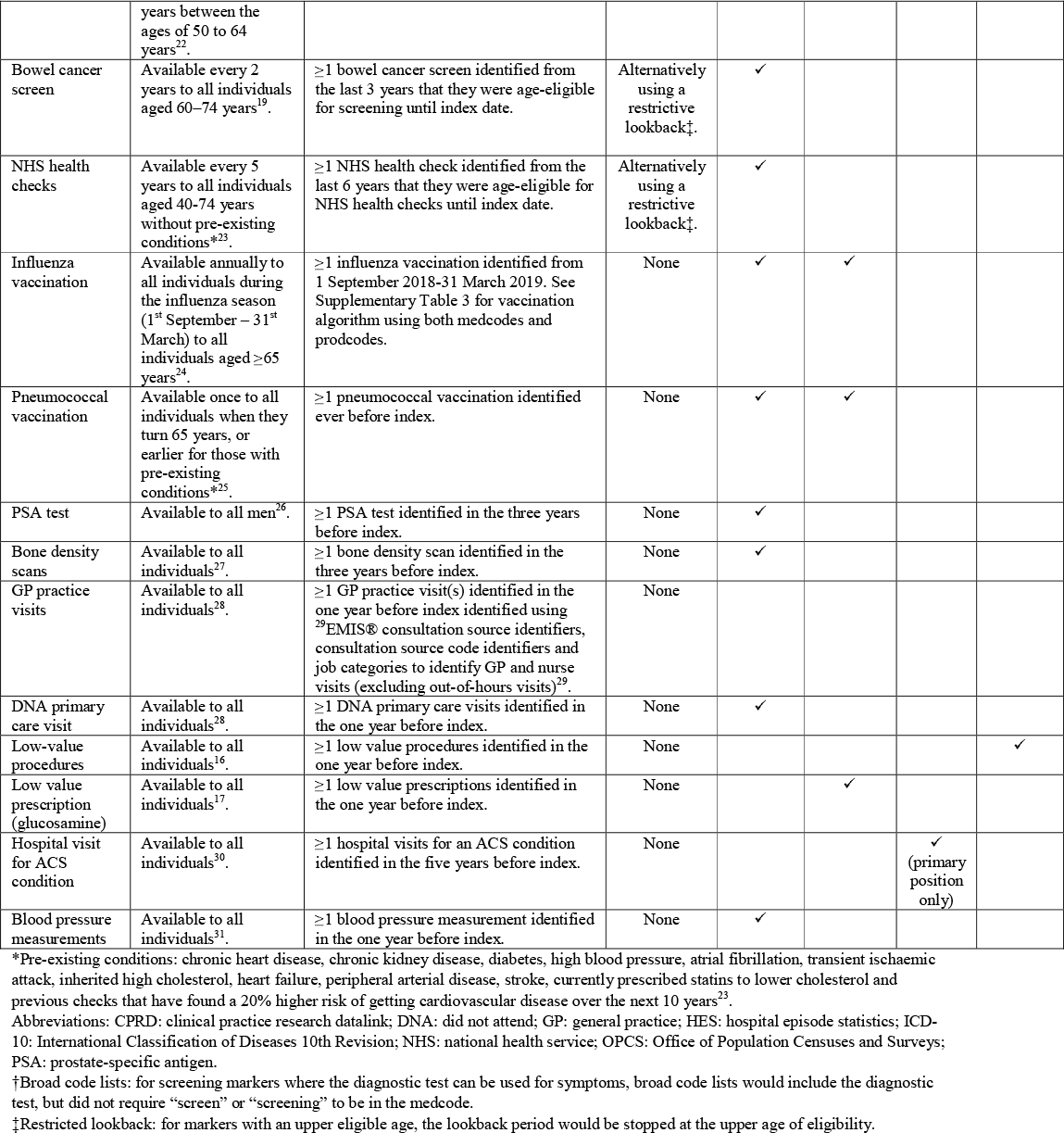
Use of markers in UK clinical practice and operational definitions.

For code lists, existing validated code lists were used where possible. Primarily we searched for code lists that were incentivised for national use through the Quality and Outcomes Framework^18^ or those that were validated through research. If codelists were not available using these sources, then they were developed using key word searches (based on Medical Subject Headings terms with corresponding synonyms). Where possible the code lists aimed to be as specific as possible (“narrow code lists”) and therefore codes were excluded if they were not clearly relevant. For example, for most screening markers we required the code to specify “screen” or “screening” but for bowel cancer we also allowed Faecal Immunochemical Tests as these are not used for symptomatic testing^19^. As a sensitivity analysis, for abdominal aortic aneurysm (AAA), breast cancer and cervical cancer screening, since the same procedure may be recorded for a screening test as for diagnostic tests investigating symptoms, we also included a broader code list that included codes that specified the relevant procedure, but did not specify “screen” or “screening”. Full inclusion and exclusion list were reviewed by a clinical epidemiologist (HIM) and differences were agreed by discussion and third-party review (EP). The search terms that were used to create the code lists and the code lists that were used can be found on LSHTM data compass (https://doi.org/10.17037/DATA.00003684).

The lookback periods for each marker were developed by firstly identifying how each of these markers are recommended for use in current UK clinical practice. For markers that are available to all at any time, the lookback period reflected the expected frequency of healthcare use in UK clinical practice. For example, for markers that were expected to be frequently recorded (e.g., blood pressure measurements) we used a one-year lookback. For markers that were expected to be less frequently recorded (e.g., hospital visit for ACS conditions) a five-year lookback was used. For markers with an upper age limit of eligibility (i.e., screening and NHS health checks), we ensured the lookback period reflected timely administration of these markers (since we were interested in capturing strong evidence of health-seeking behaviour and healthcare access). For example, breast cancer screening is offered to women every 3 years aged 50-71 years^20^ and therefore the lookback period covered the last 4-years of age-eligibility (3-years plus an additional year for uncertainty of age as only year of birth is recorded in CPRD) for breast cancer screening, until the index date. We included all follow-up time until index date to allow for delayed recording due to the transition to electronic records amongst older individuals. As a sensitivity analysis, since we were concerned that including years after the upper age of eligibility might have meant we included more symptomatic individuals rather than healthy individuals accessing screening programmes, we also employed a restricted lookback that stopped the lookback at the upper age of eligibility (see **Supplementary Figure 1**).

### Prevalence estimates

For prevalence calculations, the denominator was all individuals aged ≥66 years on 1 September 2019 and the numerator was ≥1 occurrence of the marker in the relevant lookback period. We also calculated prevalence stratified by sex (given the inclusion of several sex-specific markers) and age in 5-year bands (65-69, 70-74, 75-79, 80-84, 85-89, 90-95 and 95+ years).

We compared prevalence estimates to national estimates from PHE fingertips or from published literature, preferentially selecting for recent estimates from the UK in the relevant age group. The prevalence estimates from these sources can be found in **Table 4** and sources are detailed in **Supplementary Table 4**.

**Table 4.** Prevalence of markers.

| Variable | All Individuals | Male | Female | National estimates | Theoretical grouping from TBP model<sup>#</sup> |
| --- | --- | --- | --- | --- | --- |
| N | 1,991,284 | 915,561 | 1,075,723 |  |  |
| AAA screen* | 231,088 (11.6%) | 227,844 (24.9%) | 3,244 (0.3%) | 76.1% | Contextual |
| Breast cancer screen* | 346,116 (17.4%) | 517 (0.1%) | 345,599 (32.1%) | 71.1% | Contextual |
| Cervical cancer screen* | 397,303 (20.0%) | 153 (0.0%) | 397,150 (36.9%) | 76.2% | Contextual |
| Bowel cancer screen* | 1,439,412 (72.3%) | 687,712 (75.1%) | 751,700 (69.9%) | 60.5% | Contextual |
| NHS health checks*† | 372,244 (18.7%) | 157,484 (17.2%) | 214,760 (20.0%) | 40%‡ | Contextual |
| Influenza vaccine | 1,460,391 (73.3%) | 670,162 (73.2%) | 790,229 (73.5%) | 72.4% | Psychological |
| Pneumococcal vaccination | 1,242,359 (62.4%) | 568,798 (62.1%) | 673,561 (62.6%) | 69.0% | Psychological |
| PSA testing | 352,272 (17.7%) | 351,884 (38.4%) | 388 (0.0%) | 53.0% | Physical with active access |
| Bone density scan | 100,892 (5.1%) | 19,407 (2.1%) | 81,485 (7.6%) | 0.03-1.6% | Physical with active access |
| GP practice visits | 1,844,823 (92.6%) | 841,413 (91.9%) | 1,003,410 (93.3%) | § | Physical with active access |
| DNA primary care visit | 601,896 (30.2%) | 275,449 (30.1%) | 326,447 (30.3%) | § | Physical with lack of access |
| Low-value procedures | 358,881 (18.0%) | 168,746 (18.4%) | 190,135 (17.7%) | 0.02-0.2% | Physical with active access |
| Low-value prescription (glucosamine) | 219 (0.0%) | 75 (0.0%) | 144 (0.0%) | § | Physical with active access |
| Hospital visit for an ACS condition | 190,136 (9.5%) | 82,691 (9.0%) | 107,445 (10.0%) | 0.1% | Physical with lack of access |
| Blood pressure measurement | 1,470,006 (73.8%) | 681,294 (74.4%) | 788,712 (73.3%) | 84.6% | Physical with active access |

Abbreviations: AAA: abdominal aortic aneurysm; ACS: ambulatory care sensitive; DNA: did not attend; GP: general practice; PSA: prostate specific antigen; TPB: theory of planned behaviour.
\*The denominator for the current study does not restrict to those that are age-eligible unlike in the national estimate. For age-eligible estimates see **Figure 1**.
†The denominator for the current study does not exclude individuals without pre-existing conditions (chronic heart disease, chronic kidney disease, diabetes, high blood pressure, atrial fibrillation, transient ischaemic attack, inherited high cholesterol, heart failure, peripheral arterial disease, stroke, currently prescribed statins to lower cholesterol and previous checks that have found a 20% higher risk of getting cardiovascular disease over the next 10 years<sup>23</sup>) unlike in the national estimate.
‡The national estimate for NHS health checks is the percentage of eligible individuals receiving an NHS health check in Q1 2019/2020: 2.0%. To better match the lookback period for this marker (5-years), we multiplied this estimate by 20.
§Prevalence of these markers are not knowingly presented in national estimates. NHS digital and OpenPrescribing report the total unit counts for these markers, which are reported in **Supplementary Table 4**.

### Correlations

The correlation of all the markers within the population sample was assessed using a phi correlation matrix. The phi coefficient is designed to measure the association between binary variables, and is equivalent to a Pearson correlation when applied to binary data. It ranges from -1 to 1, where 0 signifies no relationship between the variables, 1 is a perfect positive relationship and -1 is a perfect negative relationship^32^. Variables were ordered via complete linkage hierarchical clustering which was visualised by adorning dendrograms onto the correlation matrices (heatmaply_cor in R).

The clustering of markers was compared to a theoretical grouping using the updated TBP model^5^. The theoretical grouping was based on the underlying determinants from updated TBP model (**Table 4**). Specifically, we grouped markers into four groups: those with strong psychological influences (“<u>psychologically determined</u>”; e.g., vaccinations), those with strong contextual influences (“<u>contextually determined</u>”; e.g., screening and NHS health checks) and those fully or partially dependent on physical need. Physically determined markers were further separated into those likely to represent lack of health-seeking behaviour or healthcare access (e.g., DNA primary care visit and ACS condition hospital visit; “<u>physically determined with lack of access</u>”) and those likely to represent active health-seeking behaviour or straightforward healthcare access (“<u>physically determined with active access</u>”).

All programming was conducted using R (version 4.2.1-4.2.3) and the programming code can be found on Github (https://github.com/grahams99/Health-seeking-behaviour).

## Results

Overall, 1,991,284 individuals were included (54.0% females, mean (SD) age: 75.9 (7.4); **Supplementary Table 5**).

The prevalence of markers in the overall population ranged from <0.1% for low value prescriptions to 92.6% for GP visits. The proportion with at least one GP visit was so high that we conducted a *post-hoc analysis* that revealed the median (IQR) number of GP visits was 7 (4-11) with some patients having over 25 visits per year (**Supplementary Figure 2**). The prevalence of markers was similar between males and females, except for sex-specific markers (**Table 4**). For screening and NHS health checks, broad code lists with standard lookback periods had the highest prevalence, whereas narrow code list with restrictive lookback had the lowest. For AAA screening and NHS health checks, changing the operational definition changed the prevalence <2% (**Supplementary Table 6**). The prevalence of most markers was in line with national estimates, particularly for the vaccinations, PSA testing and bone density scans. For example, 73.3% individuals in the current study had an influenza vaccination with national estimates reporting 72.4% influenza vaccination uptake among ≥65-year-olds in the 2019/20 influenza vaccination season^33^. The prevalence of screening and NHS health checks in the overall population was lower than national estimates, although this generally improved with comparison to currently eligible age-groups (**Figure 1**). Hospital visit for an ACS condition were higher than literature estimates as it was not possible to differentiate planned and unplanned hospitalisations in the current datasets (9.5% in current study vs. 0.1% in literature).

**Figure 1.**
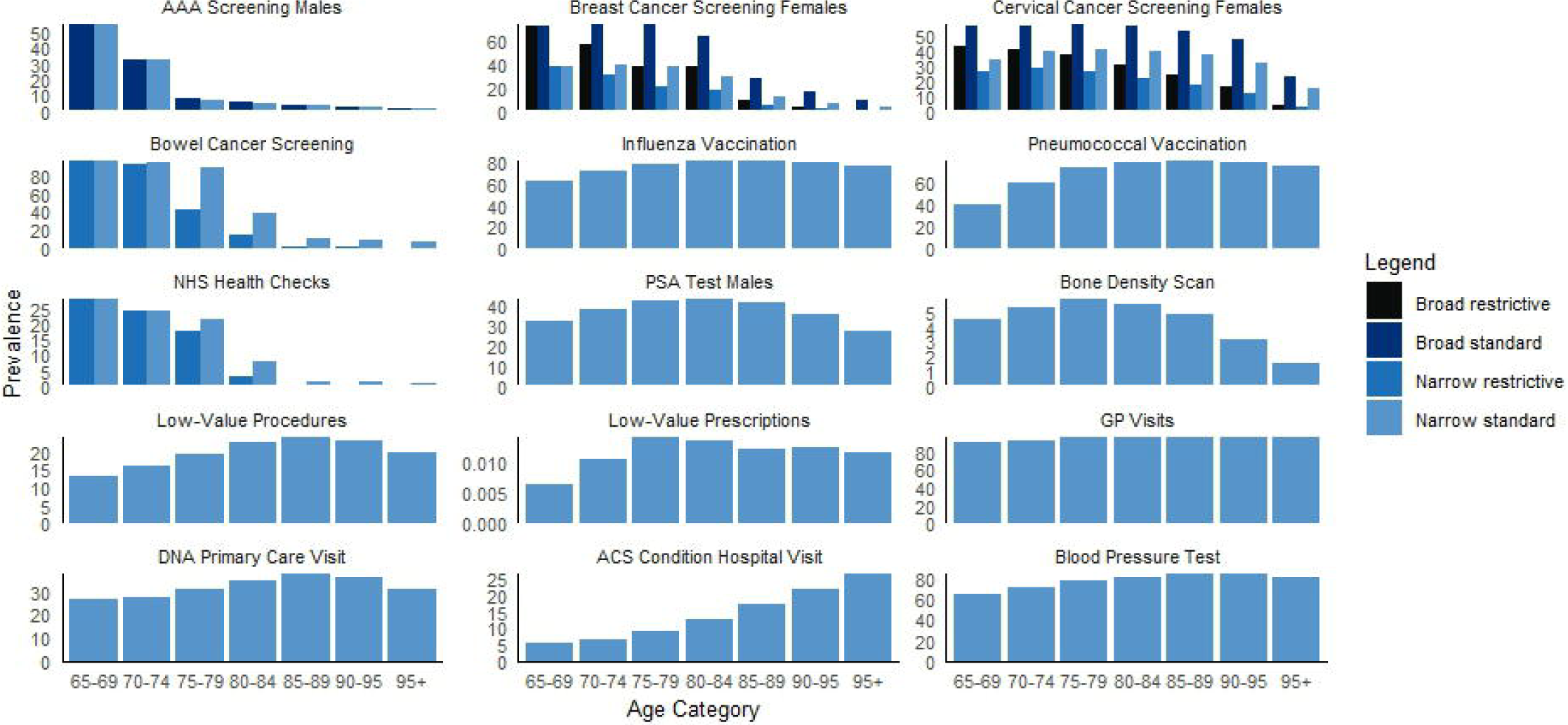
Prevalence of markers, stratified by age category. Abbreviations: AAA: abdominal aortic aneurysm; DNA: do not attend; GP: general practice; NHS: National Health Service. Note: the numbers and proportions for these bar charts can be found in Supplementary Table 7.

The prevalence of markers typically varied by age category, with a number of patterns evident (**Figure 1**). The recorded prevalence of markers with upper age eligibility (screening and NHS health checks) decreased with age (e.g., 28.0% in 65–69 year-olds vs 1.5% in 85–89 year-olds for NHS health checks), whereas the prevalence of ACS conditions, blood pressure measurements and vaccinations rose with age (e.g. 62.9% in 65–69 year-olds vs 80.5% in 85–89 year-olds for influenza vaccination). Although more common in younger age groups, screening marker prevalence still fell short of national estimates in currently eligible age-groups (e.g., 54.3% in 65–69 year-olds vs 76.1% in national estimates for AAA screening in men). PSA tests, bone density scans, low-value procedures, low-value prescriptions and DNA primary care visits peaked at 75-89 years, with lower prevalence in younger and older individuals. GP visits were consistent across age categories. As expected, the proportion of individuals with ≥1 GP practice visit was very high. The *post-hoc analysis* revealed the number of GP visits increased by age category until the last age strata (90+ years), when it decreased slightly (**Supplementary Figure 3**).

Using broad rather than narrow code lists, the estimated prevalences were similar for AAA screening across all age strata and for breast cancer screening in those aged 65-69 years. For all other breast cancer screening strata and for cervical cancer screening, broad code lists resulted in a higher prevalence than narrow. For standard versus restricted lookback periods, the prevalence was the same for individuals entering the cohort below the upper age of eligibility of that marker, whereas after this point there was lower prevalence in the restricted versus standard age strata.

In the overall study population, unsurprisingly, GP visits were strongly correlated with blood pressure measurements (phi φ 0.42) and influenza vaccination (0.33). Blood pressure measurements were also strongly correlated with influenza vaccination (0.23). Markers with the strongest negative correlation were blood pressure measurements and NHS health checks (-0.14) (**Figure 2**). Among males, GP visits and blood pressure measurements had the strongest positive correlation (0.45), followed by influenza and pneumococcal vaccinations (0.42). Other strong correlations included GP visits and pneumococcal vaccination (0.36) and blood pressure measurements and influenza vaccination (0.25) (**Figure 2**). Among females, GP visits were also strongly correlated with blood pressure measurements (0.40) and blood pressure measurements with influenza vaccination (0.30). There were also strong correlations between pneumococcal vaccination with influenza vaccination (0.39), bowel cancer screening and NHS health checks (0.23) (**Figure 2**).

**Figure 2.**
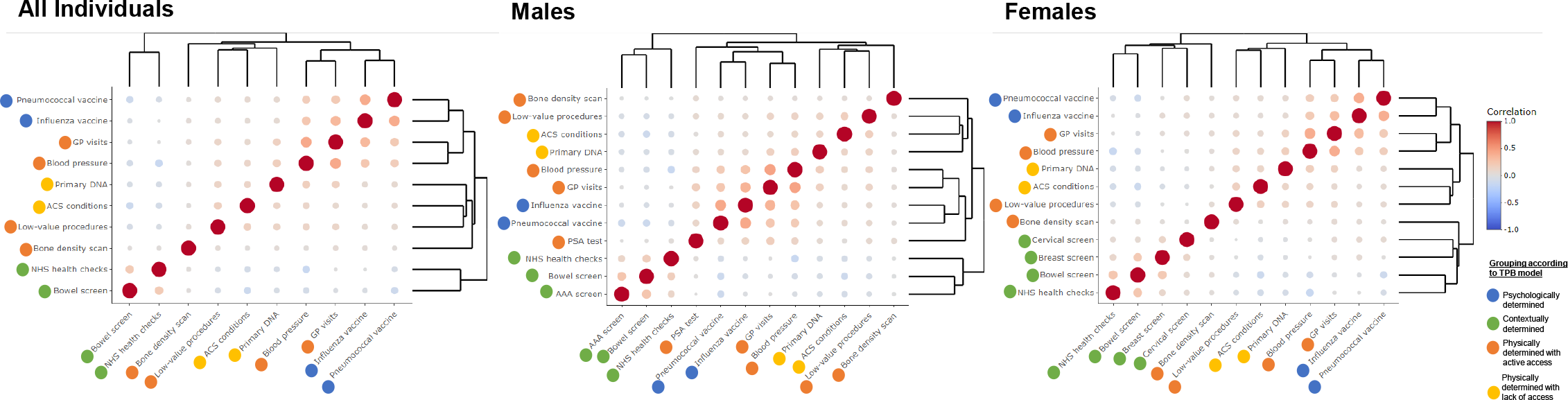
Correlation matrix plots. Abbreviations: ACS: ambulatory care sensitive; DNA: did not attend; NHS: National Health Service. The correlations are calculated using phi coefficient for binary variables. The clustering is visualised through the adorned dendrograms which are ordered via complete linkage hierarchical clustering. The size and the shading of the bubble represents the strength of the correlation. Note: the correlation coefficients for these plots can be found in **Supplementary Table 8-10**.

Markers that were clustered together in the correlation matrices were: 1) blood pressure measurements, GP visits and influenza and pneumococcal vaccinations; 2) NHS health checks and bowel, cervical, breast cancer and AAA screening; and 3) ACS conditions, primary care DNA, bone density scans and low-value procedures. Markers from group 2) generally had a weak negative correlation with markers from group 3). When comparing these data-driven clusters with the theoretical grouping of markers there were some similarities. In both methods the “contextually determined” (i.e., NHS health checks and screenings) were grouped together as well as the “physically determined with a lack of healthcare access” (i.e., ACS conditions and primary care DNA). On the other hand, GP visits and blood pressure measurements were grouped with “psychologically determined” markers in the data-driven approach, but with the “physically determined with active healthcare access” in the theoretical grouping.

## Discussion

### A statement of the principal findings

Overall, this study found that it is feasible to identify markers of health-seeking behaviour and healthcare access in UK EHRs. The prevalence of these markers ranged significantly and were generally in line with national estimates. Screening and NHS health checks were under-recorded in the EHR data, although prevalence was closer to national estimates amongst younger age groups that were currently eligible for these programmes. The prevalence and pattern of markers differed by age, with AAA screening declining with older age and hospital visits for ACS condition increasing. Correlations between markers revealed clusters that aligned well with theoretical groupings informed by the updated TBP model based on psychological, contextual and physical underlying determinants.

### Strengths and weaknesses of the study

To our knowledge, this is the first study that has systematically identified proxies or markers of health-seeking behaviour or healthcare access using routinely collected data in the UK. Previous studies have adjusted for variables which may reflect confounding by health-seeking behaviour such as GP consultations, but without an explicit framework for selecting these. Our study demonstrates that a framework is beneficial since health-seeking behaviour and healthcare access are complex phenomena with multiple determinants, which may behave differently, and vary by age and sex. Linkage across primary and secondary care also strengthened this study as different types of healthcare utilisation with different underlying determinants could be captured. We included a large and representative cohort of over 2 million individuals aged 66 years and over in England. For some of the markers, older individuals might not have been historically eligible for services, which represents an important caveat during the interpretation of prevalence estimates. However, we accounted for this by calculating age-stratified prevalence. The study also only measured markers of health-seeking behaviour and healthcare access at a single point in time: these characteristics are not static, and individual behaviour and service accessibility can change over time. In addition, for some of the identified markers the influence of health need could not be entirely separated from health-seeking behaviour and healthcare access and therefore in some cases prevalence would be driven to some extent by health need. These findings might also not be generalisable to younger individuals where perhaps there are other contextual determinants to consider (e.g., occupation)^9^.

### Strengths and weaknesses in relation to other studies / discussing important differences in results

Previous studies that have used EHR to identify markers of health-seeking behaviour and healthcare access in the US^6-8^ are in a considerably different context from the UK in terms of the healthcare system, claims-based recording systems and underlying determinants of health. This is likely to explain the different prevalence of markers identified in the current study. For example, the prevalence of pneumococcal vaccination was only around 11.4% in a study of ≥65 year olds identified in the Medicare database with an influenza vaccination during the 2019/2020 season^7^, whereas the prevalence was 62.4% in the current study. These differences support the importance of context-specific markers of health-seeking behaviour and healthcare access.

Our study adds to a growing body of literature highlighting the potential to capture proxies of healthcare access and health-seeking behaviour. In prior studies in the US, these proxies were included as confounders during estimation of vaccine effectiveness, and they could play a similar role during observational studies in a UK context.^7^

### The meaning of the study: possible explanations and implications for researchers, clinicians and policymakers

Based on the findings presented here, we propose several recommendations and considerations for researchers that wish to identify health-seeking behaviour and healthcare access in EHRs – whether to study healthcare use directly, or to quantify or adjust for confounding.

First, a range of different markers are required to fully represent both active health-seeking behaviour and healthcare access, or lack of these. Since health-seeking behaviour/healthcare access is such a complex phenomenon, it may be useful to include markers with different underlying determinants from the updated TBP model (psychologically, contextually and physically determined). If multiple markers are available, they can be included as separate confounders in multivariate models, or researchers may wish to consider tools such as high-dimensional propensity scores to guide study-specific confounder identification, prioritisation and adjustment^34^.

Second, the optimal code lists will depend on the precise research question. Narrow code lists (e.g., using government incentivised code lists) can identify markers of health-seeking behaviour and healthcare access with high specificity. Broader code lists will capture more events, but may be more influenced by underlying health need. For markers with specific age-eligibility (e.g., screening or NHS health checks) look-back periods that restrict to time periods when individuals were age eligible improved specificity. However, more relaxed lookback periods might be preferred if there are expected to be artefacts in data recording such as transfer of historical information to electronic health records.

Third, prior to adjusting for health-seeking behaviour and healthcare access, interactions by age, sex and underlying health conditions should be considered. Markers that were recently introduced into clinical practice (e.g., AAA screening was introduced in the UK in 2013^35^) will likely decrease in prevalence with increasing age and can be supplemented with markers that increase with increasing age (e.g., ACS conditions). Otherwise, markers with relatively consistent prevalence across age strata are available (e.g., GP visits or blood pressure measurements). If markers that are restricted to specific sex (e.g., breast cancer screening) are utilised then these can be supplemented with markers of the opposite sex (e.g., AAA screening). For markers where there is some partial influence of underlying health conditions (e.g., pneumococcal vaccinations recommended to all but may be more highly prioritised among those with high-risk conditions) can be supplemented with markers that are administered to those that are healthier (e.g., NHS health checks).

### Unanswered questions and future research

Future researchers who are concerned with potential confounding from health-seeking behaviour and healthcare access in their study can use these markers to quantify and adjust for confounding. Where possible, a range of markers with different underlying determinants from the updated TPB model should be used and possible interactions by age, sex and underlying condition should be considered. Future research may identify key confounders within each theoretical group or cluster that are sufficient for confounding adjustment, although these are likely to be study-specific.

Common data models across datasets could increase efficiency and comparability of research investigating or adjusting for health-seeking behaviour and healthcare access, but future research is needed to identify suitable markers in alternative datasets and establish comparability. Additional markers may be identified in alternative datasets using the developed criteria.

## Conclusion

Overall, markers of health-seeking behaviour and healthcare access can be identified in UK EHR, with prevalence estimates in line with national estimates. National screening programme estimates still fell short of national estimates even when restricting to currently eligible age groups. The generally low correlations between different proxy markers of health-seeking behaviour and healthcare access, and different age-profiles of markers, suggest a range of variables are needed to capture different determinants of healthcare use.

## Data Availability

These data were obtained from the Clinical Practice Research Datalink, provided by the UK Medicines and Healthcare products Regulatory Agency. The authors' licence for using these data does not allow sharing of raw data with third parties. Information about access to Clinical Practice Research Datalink data is available here: https://www.cprd.com/research-applications. Codelists for this study are available at https://doi.org/10.17037/DATA.00003684 and code at https://github.com/grahams99/Health-seeking-behaviour.

https://github.com/grahams99/Health-seeking-behaviour

## Contributors

Study concepts: SG, NA, JLW and HIM; study design: all authors; data acquisition: SG; Programming: SG, EPKP and HIM; Statistical analysis: SG; Supervision: EPKP, NA, JLW and HIM; Interpretation of results: all authors; Manuscript preparation: SG; Manuscript editing: all authors; manuscript review: all authors; Manuscript approval: all authors.

## Disclaimer

The views expressed are those of the authors and not necessarily those of the NHS, the NIHR, or UKHSA.

## Patient consent for publication

Not required.

## Ethics approval

The protocol for the study received scientific and ethical approval from the CPRD Research Data Governance committee (#22_002202).

## Provenance and peer review

Not commissioned; externally peer reviewed.

## Patient involvement statement

No patient or public involvement in the study as anonymised patient dataset was used in the analysis.

