## Supplementary information for "Identifying markers of health-seeking behaviour and healthcare access in UK electronic health records"

### Tables

#### Supplementary Table 1. Current available literature and markers derived from each

| Author and year | Study type | Markers identified | Included/excluded | Reason for exclusion |
|---|---|---|---|---|
| Izurieta et al, 2020[1] | Observational cohort study of relative influenza vaccine effectiveness using US claims data | Pneumococcal vaccination | Included | |
| | | Annual wellness visit | Included as NHS health checks | |
| | | Bone mass measurements | Included as bone density scan | |
| | | Cardiovascular disease screen tests | Excluded | Failed criteria 3 |
| | | Colorectal cancer screen | Included as bowel cancer screen | |
| | | Diabetes screen | Included as NHS health checks | |
| | | Initial Preventive Physical Examination | Included as NHS health checks | |
| | | Prostate Cancer Screen | Included as PSA test | |
| | | Screen Mammography | Included as breast cancer screen | |
| | | Screen Pap test | Included as cervical cancer screen | |
| | | Screen pelvic examination | Excluded | Failed criteria 3 |
| | | Depression screen | Excluded | Failed criteria 3 |
| | | Other preventative services | Excluded | Unclear what this entails |
| Izurieta et al, 2021[2] | Observational cohort study of influenza vaccine effectiveness using US claims data | Annual Wellness Visits | Included as NHS health checks | |
| | | Counseling & Health Risk Assessment | Excluded | Failed criteria 3 |
| | | Pneumococcal Vaccination | Included | |
| | | Tetanus-containing vaccination | Included | Failed criteria 3 |
| | | Shingrix vaccination | Excluded | Failed criteria 1 |
| Zhang et al, 2017[3] | Observational cohort study of influenza vaccine effectiveness using US claims data | Outpatient visits | Included as GP visits | |
| | | Hospital visits | Excluded | Failed criteria 3 |
| | | Colonoscopies | Included as bowel cancer screen | |
| | | Fecal occult blood tests | Included as bowel cancer screen | |

Abbreviations: DNA: did not attend; GP: general practice; NHS: national health service; PSA: prostate-specific antigen.

#### Supplementary Table 2. How each of the markers meet each of the criteria

| Marker | **Criteria 1** Should have been currently or recently available in national clinical practice to all individuals (overall or by sex) at cohort entry. | **Criteria 2** Should be routinely recorded in the available data sources. | **Criteria 3** Should not be primarily dependent on underlying health needs. |
|---|---|---|---|
| AAA screen | ~<br><br>AAA screening was introduced in 2013 in the UK[4] so for men entering the cohort at age 71 years and above they might not have ever been offered a screen (since this is offered to all men when they turn 65 years). | ✓ | ✓<br><br>Universally available except for men that have been treated for an AAA previously. The incidence of AAA in those under 65 years is very low[5]. |
| Breast cancer screen | ✓ | ✓ | ✓ |

| **Marker** | **Criteria 1**<br>Should have been currently or recently available in national clinical practice to all individuals (overall or by sex) at cohort entry. | **Criteria 2**<br>Should be routinely recorded in the available data sources. | **Criteria 3**<br>Should not be primarily dependent on underlying health needs. |
|---|---|---|---|
| Cervical cancer screen | ✓ | ✓ | ✓ |
| Bowel cancer screen | ✓ | ✓ | ✓ |
| NHS health checks | ✓ | ✓ | ~<br>Universally available to those without pre-existing high-risk conditions[6]. |
| Influenza vaccination | ✓ | ✓ | ✓ |
| Pneumococcal vaccination | ✓ | ✓ | ✓ |
| PSA testing | ✓ | ✓ | ~<br>Men can request a PSA test with no prior diagnosis of prostate cancer who are asymptomatic, men presenting with symptoms, and men who have had a previous high PSA level and are being monitored[7]. |
| Bone density scans | ✓ | ✓ | ~<br>Can be requested for individuals over 50 years with a risk of developing osteoporosis or for those with other risk factors such as smoking or broken bone[8]. |
| GP practice visits | ✓ | ✓ | ~<br>Occurs for those with symptoms. |
| Low-value procedures | ✓ | ✓ | ~<br>Requested for those with symptoms. |
| Low-value prescriptions | ✓ | ~<br>Although these can be prescribed in primary care, individuals can also buy some low values prescriptions over the counter and therefore we might see under recording of this. However, those that access primary care for something that can be bought over the counter likely have very active access to healthcare. | ✓ |
| Hospital visit for an ACS condition | ✓ | ✓ | ~<br>Occurs for those with symptoms. |
| DNA primary care visit | ✓ | ✓ | ~<br>Occurs for those with symptoms. |
| Blood pressure measurements | ✓ | ~ | ~<br>Universally available, but disproportionately requested for those with underlying health conditions. |

Abbreviations: DNA: did not attend; GP: general practice; NHS: national health service; PSA: prostate-specific antigen.

### Supplementary Table 3. Influenza Vaccination Algorithm

| Combination of codes on same day | Total number of vaccination events | Decision | Rationale |
|---|---|---|---|
| Given and neutral | 725432 | Record as valid vaccination. | |
| Given and absent | 1149 (of these only 924 are the first vaccination dose) | Do not record as valid vaccination. | The prevalence of this marker may be underestimated very slightly, however, in this instance better to be more specific than sensitive when it comes to confounders[9]. |
| Given and adverse | 14 | Record as valid vaccination. | Likely that this patient received the vaccination, but then had an adverse event on the same day. |
| Given and product | 395881 | Record as valid vaccination. | |
| Given and given with lag | 4700 | Record as valid vaccination. | |
| Neutral and absent | 1178 | Do not record as valid vaccination. | |
| Neutral and adverse | 9 | Record as valid vaccination. | |
| Neutral and product | 295746 | Record as valid vaccination. | |
| Neutral and given lag | 7748 | Record as valid vaccination. | |
| Absent and adverse | 27 | Do not record as valid vaccination. | Likely that these patients are reporting a previous adverse event as the reason for not wanting to get vaccinated. |
| Absent and product | 218 (of these only 194 are first vaccination dose) | Do not record as valid vaccination. | The prevalence of this marker will be underestimated very slightly, however, in this instance better to be more specific than sensitive when it comes to confounders[9]. |
| Absent and given with lag | 570 | Record as valid vaccination. | This most likely reflects that the reason a vaccine wasn't given on the event date is that the patient had already had it elsewhere. We can be reasonably confident that the patient was vaccinated, but we don't know the exact date. |
| Adverse and product | 1 | Record as valid vaccination. | Likely that this patient received the vaccination, but then had an adverse event on the same day. |
| Adverse and given with lag | 0 | Ignore – no events. | |
| Given with lag and product | 784 | Record as valid vaccination. | |

Note: since influenza vaccinations can be identified using both medcodes and prodcodes and since medcodes do not always insinuate presence of a vaccination, an algorithm was developed for combinations of codes that occurred on the same day. Medcodes were separated into those that were clearly given ("given" or "administered"), given with delay ("given" or "administered" but evidence this occurred in another setting previously), neutral (vaccination mentioned but no "given" or "administered") and absent (vaccination "refused" or "not consented). Then we looked at vaccination events (using both medcodes and prodcodes) that were recorded on the same date and categorised these according to the above framework.

### Supplementary Table 4. National estimates

| Marker | Estimate (%, unless otherwise specified) | Year | Numerator | Denominator |
|---|---|---|---|---|
| AAA screen[10] | 76.1 | 2019/20 | Number of men eligible for the initial screen who have had a conclusive scan result within the screening year plus an additional 3 months (in the event of non attendance and cancellations at the end of the year this allows men to be reinvited and screened). | Number of eligible men in their 65th year to whom the screening programme propose that a screening encounter during the reporting period should be offered. |
| Breast cancer screen[11] | 71.1 | 2019/20 | The number of persons registered to the practice who were screened adequately in the previous 36 months | The number of eligible persons on last day of the review period |
| Cervical cancer screen[12] | 76.2 | 2019 | The number of women in the resident population eligible for cervical screening aged 50 to 64 years at end of period reported who were screened adequately within the previous 5.5 years. | The number of women in the resident population eligible for cervical screening aged 50 to 64 years |
| Bowel cancer screen[13] | 60.5 | 2019 | Adequately screened (numerator) is the number of eligible men and women who have had an adequate gFOBT screening result recorded in the past 30 months. | Eligible population (denominator) is the number of men and women aged 60 to 74 years resident in the area (determined by postcode of residence) who are eligible for bowel cancer screening at a given point in time, excluding those whose recall has been ceased for clinical reasons (e.g. no functioning colon) or if they opt out of the programme. |

| | | | | |
|---|---|---|---|---|
| NHS health checks[14] | 2.0 | 2019/20 Q1 | Number of people aged 40-74 eligible for an NHS Health Check who were recorded as receiving an NHS Health Check in the current quarter | Number of people aged 40-74 eligible for an NHS Health Check in the financial year. |
| Influenza vaccination[15] | 72.4 | 2019/20 | The number of adults aged 65 and over, who received the flu vaccination between 1st September to the end of February as recorded in the GP record. | Number of adults aged 65 years and older |
| Pneumococcal vaccination[16] | 69 | 2019/20 | These data describe pneumococcal polysaccharide vaccine (PPV) uptake for the survey year, for those aged 65 years and over. | Those aged 65 years and over |
| PSA test[17] | 52.95 | 2002-2011 | Number of men with at least one PSA test identified over a 10 year period | Number of men |
| Bone density scan[18] | 0.3 to 16.2 per 1000 weighted population | 2013/14 | Number of bone scans. | Total population. |
| GP practice visits[19] | 39127 per 100 000 patient-months | 2019 | Number of primary care consultations in CPRD Aurum | Number of patient months |
| GP practice visits[19] | 26 919 consultations per 100 000 patient-months | 2020 | Number of primary care consultations in CPRD Aurum | Number of patient months |
| DNA primary care visit[19] | Number: 23578484 | Dec-19 | Number of primary care appointments that were DNA | |
| Low value procedures[20] | 0.02-0.2 | 2017/18 | Number of category 1 interventions. | Total English population, age and sex-standardised. |
| Low-value prescriptions (glucosamine) | 21,961 items | 2019/20 | Total quantity prescribed in primary care | |
| Hospital visit for an ACS condition[21] | 0.1 | 2018/19 | Number of unplanned hospitalisations for chronic ambulatory care sensitive conditions | Total unplanned hospitalisations |
| Blood pressure test[22] | 84.6 | 2012-13 | Number of individuals aged ≥50 years with a blood pressure check in the last year | Number of individuals aged ≥50 years |

Abbreviations: DNA: did not attend; GP: general practice; NHS: national health service; PSA: prostate-specific antigen.

### Supplementary Table 5. Study population.

| Group | Category | All N= 1,991,284 |
|---|---|---|
| Age continuous, mean (SD) | | 75.9 (7.4) |
| Age category, N (%) | 65-69 | 448,063 (22.5%) |
| | 70-74 | 557,599 (28.0%) |
| | 75-79 | 401,290 (20.2%) |
| | 80-84 | 295,492 (14.8%) |
| | 85-89 | 181,835 (9.1%) |
| | 90-95 | 80,943 (4.1%) |
| | 95+ | 26,062 (1.3%) |
| Sex, N (%) | Female | 1,075,723 (54.0%) |
| Ethnicity, N (%) | Asian | 67,961 (3.4%) |
| | Black | 36,912 (1.9%) |
| | Mixed | 10,072 (0.5%) |
| | Other | 17,711 (0.9%) |
| | White | 1,759,754 (88.4%) |
| | Missing | 98,874 (5.0%) |
| Region, N (%) | East Midlands | 42,106 (2.1%) |
| | East of England | 95,413 (4.8%) |
| | London | 263,825 (13.2%) |
| | North East | 67,278 (3.4%) |

| | | |
|---|---|---|
| | North West | 381,592 (19.2%) |
| | South East | 438,407 (22.0%) |
| | South West | 270,484 (13.6%) |
| | West Midlands | 357,363 (17.9%) |
| | Yorkshire and The Humber | 74,805 (3.8%) |
| | Unknown | 11 (0.0%) |

Abbreviations: N: number; SD: standard deviation.

### Supplementary Table 6. Prevalence of markers using different definitions

| **Variable** | **All** | **Male** | **Female** |
|---|---|---|---|
| N | 1,991,284 | 915,561 | 1,075,723 |
| AAA screen | 231,088 (11.6%) | 227,844 (24.9%) | 3,244 (0.3%) |
| AAA screen broad* | 238,186 (12.0%) | 233,574 (25.5%) | 4,612 (0.4%) |
| Breast cancer screen restrictive† | 253,610 (12.7%) | 227 (0.0%) | 253,383 (23.6%) |
| Breast cancer screen | 346,116 (17.4%) | 517 (0.1%) | 345,599 (32.1%) |
| Breast cancer screen broad* restrictive† | 484,402 (24.3%) | 573 (0.1%) | 483,829 (45.0%) |
| Breast cancer screen broad* | 686,521 (34.5%) | 1,517 (0.2%) | 685,004 (63.7%) |
| Cervical cancer screen restrictive† | 256,565 (12.9%) | 40 (0.0%) | 256,525 (23.8%) |
| Cervical cancer screen | 397,303 (20.0%) | 153 (0.0%) | 397,150 (36.9%) |
| Cervical cancer screen broad* restrictive† | 373,369 (18.8%) | 93 (0.0%) | 373,276 (34.7%) |
| Cervical cancer screen broad* | 588,288 (29.5%) | 361 (0.0%) | 587,927 (54.7%) |
| NHS health checks restrictive† | 338,441 (17.0%) | 143,325 (15.7%) | 195,116 (18.1%) |
| NHS health checks | 372,244 (18.7%) | 157,484 (17.2%) | 214,760 (20.0%) |
| Bowel screen restrictive† | 1,151,943 (57.8%) | 553,109 (60.4%) | 598,834 (55.7%) |
| Bowel screen | 1,439,412 (72.3%) | 687,712 (75.1%) | 751,700 (69.9%) |

Abbreviations: AAA: abdominal aortic aneurysm; DNA: did not attend; GP: general practice; PSA: prostate-specific antigen.
*Broad code list: code lists were less specific e.g., screening markers could mention the relevant test, but without requiring “screen” in the code.
†Restrictive lookback: for markers with an upper age of eligibility (e.g., cancer screening and NHS health checks) the lookback period stopped at the age of upper eligibility.

### Supplementary Table 7. Prevalence of markers by age.

| Marker | Age category | Count | Prevalence |
|---|---|---|---|
| AAA screen broad* | 65-69 | 121,345 | 55.0 |
| AAA screen broad* | 70-74 | 87,776 | 32.7 |
| AAA screen broad* | 75-79 | 13,957 | 7.4 |
| AAA screen broad* | 80-84 | 6,930 | 5.3 |
| AAA screen broad* | 85-89 | 2,748 | 3.7 |
| AAA screen broad* | 90-95 | 709 | 2.5 |
| AAA screen broad* | 95+ | 109 | 1.6 |
| AAA screen | 65-69 | 119,707 | 54.3 |
| AAA screen | 70-74 | 85,529 | 31.8 |
| AAA screen | 75-79 | 13,075 | 7.0 |
| AAA screen | 80-84 | 6,360 | 4.9 |
| AAA screen | 85-89 | 2,445 | 3.3 |
| AAA screen | 90-95 | 634 | 2.3 |
| AAA screen | 95+ | 94 | 1.4 |
| Breast cancer screen broad* | 65-69 | 165,587 | 72.8 |
| Breast cancer screen broad* | 70-74 | 214,147 | 74.1 |
| Breast cancer screen broad* | 75-79 | 160,055 | 74.9 |
| Breast cancer screen broad* | 80-84 | 104,683 | 63.5 |
| Breast cancer screen broad* | 85-89 | 30,543 | 28.2 |
| Breast cancer screen broad* | 90-95 | 8,351 | 15.8 |
| Breast cancer screen broad* | 95+ | 1,638 | 8.5 |
| Breast cancer screen broad* restrictive† | 65-69 | 165,575 | 72.8 |
| Breast cancer screen broad* restrictive† | 70-74 | 163,148 | 56.4 |
| Breast cancer screen broad* restrictive† | 75-79 | 81,247 | 38 |
| Breast cancer screen broad* restrictive† | 80-84 | 62,039 | 37.6 |
| Breast cancer screen broad* restrictive† | 85-89 | 10,162 | 9.4 |
| Breast cancer screen broad* restrictive† | 90-95 | 1,500 | 2.8 |
| Breast cancer screen broad* restrictive† | 95+ | 158 | 0.8 |
| Breast cancer screen | 65-69 | 86,335 | 37.9 |
| Breast cancer screen | 70-74 | 112,561 | 38.9 |
| Breast cancer screen | 75-79 | 81,241 | 38 |
| Breast cancer screen | 80-84 | 48,422 | 29.4 |
| Breast cancer screen | 85-89 | 13,182 | 12.2 |
| Breast cancer screen | 90-95 | 3,300 | 6.2 |
| Breast cancer screen | 95+ | 558 | 2.9 |
| Breast cancer screen restrictive† | 65-69 | 86,327 | 37.9 |
| Breast cancer screen restrictive† | 70-74 | 87,674 | 30.3 |
| Breast cancer screen restrictive† | 75-79 | 44,831 | 21 |
| Breast cancer screen restrictive† | 80-84 | 29,008 | 17.6 |
| Breast cancer screen restrictive† | 85-89 | 4,817 | 4.4 |
| Breast cancer screen restrictive† | 90-95 | 679 | 1.3 |
| Breast cancer screen restrictive† | 95+ | 47 | 0.2 |
| Cervical cancer screen broad* | 65-69 | 127,792 | 56.2 |

| | | | |
|---|---|---|---|
| Cervical cancer screen broad* | 70-74 | 160,051 | 55.4 |
| Cervical cancer screen broad* | 75-79 | 122,449 | 57.3 |
| Cervical cancer screen broad* | 80-84 | 91,696 | 55.6 |
| Cervical cancer screen broad* | 85-89 | 57,058 | 52.7 |
| Cervical cancer screen broad* | 90-95 | 24,576 | 46.5 |
| Cervical cancer screen broad* | 95+ | 4,305 | 22.3 |
| Cervical cancer screen broad* restrictive† | 65-69 | 95,951 | 42.2 |
| Cervical cancer screen broad* restrictive† | 70-74 | 114,760 | 39.7 |
| Cervical cancer screen broad* restrictive† | 75-79 | 78,703 | 36.8 |
| Cervical cancer screen broad* restrictive† | 80-84 | 49,255 | 29.9 |
| Cervical cancer screen broad* restrictive† | 85-89 | 25,412 | 23.5 |
| Cervical cancer screen broad* restrictive† | 90-95 | 8,537 | 16.1 |
| Cervical cancer screen broad* restrictive† | 95+ | 658 | 3.4 |
| Cervical cancer screen | 65-69 | 76,059 | 33.4 |
| Cervical cancer screen | 70-74 | 112,676 | 39 |
| Cervical cancer screen | 75-79 | 85,484 | 40 |
| Cervical cancer screen | 80-84 | 63,964 | 38.8 |
| Cervical cancer screen | 85-89 | 39,581 | 36.6 |
| Cervical cancer screen | 90-95 | 16,649 | 31.5 |
| Cervical cancer screen | 95+ | 2,737 | 14.2 |
| Cervical cancer screen restrictive† | 65-69 | 59,641 | 26.2 |
| Cervical cancer screen restrictive† | 70-74 | 81,978 | 28.4 |
| Cervical cancer screen restrictive† | 75-79 | 55,434 | 25.9 |
| Cervical cancer screen restrictive† | 80-84 | 35,276 | 21.4 |
| Cervical cancer screen restrictive† | 85-89 | 17,966 | 16.6 |
| Cervical cancer screen restrictive† | 90-95 | 5,846 | 11.1 |
| Cervical cancer screen restrictive† | 95+ | 384 | 2.0 |
| Bowel cancer screen | 65-69 | 423,248 | 94.5 |
| Bowel cancer screen | 70-74 | 521,588 | 93.5 |
| Bowel cancer screen | 75-79 | 354,378 | 88.3 |
| Bowel cancer screen | 80-84 | 114,222 | 38.7 |
| Bowel cancer screen | 85-89 | 17,514 | 9.6 |
| Bowel cancer screen | 90-95 | 6,743 | 8.3 |
| Bowel cancer screen | 95+ | 1,719 | 6.6 |
| Bowel cancer screen restrictive† | 65-69 | 423,248 | 94.5 |
| Bowel cancer screen restrictive† | 70-74 | 513,796 | 92.1 |
| Bowel cancer screen restrictive† | 75-79 | 170,462 | 42.5 |
| Bowel cancer screen restrictive† | 80-84 | 42,108 | 14.3 |
| Bowel cancer screen restrictive† | 85-89 | 1,903 | 1.0 |
| Bowel cancer screen restrictive† | 90-95 | 391 | 0.5 |
| Bowel cancer screen restrictive† | 95+ | 35 | 0.1 |
| NHS health checks | 65-69 | 125,225 | 27.9 |
| NHS health checks | 70-74 | 135,076 | 24.2 |
| NHS health checks | 75-79 | 85,472 | 21.3 |
| NHS health checks | 80-84 | 22,626 | 7.7 |
| NHS health checks | 85-89 | 2,781 | 1.5 |

| | | | |
|---|---|---|---|
| NHS health checks | 90-95 | 844 | 1.0 |
| NHS health checks | 95+ | 220 | 0.8 |
| NHS health checks restrictive† | 65-69 | 125,225 | 27.9 |
| NHS health checks restrictive† | 70-74 | 134,622 | 24.1 |
| NHS health checks restrictive† | 75-79 | 69,649 | 17.4 |
| NHS health checks restrictive† | 80-84 | 8,933 | 3.0 |
| NHS health checks restrictive† | 85-89 | 6 | <0.1 |
| NHS health checks restrictive† | 90-95 | 6 | <0.1 |
| NHS health checks restrictive† | 95+ | 0 | 0 |
| Influenza vaccination | 65-69 | 281,993 | 62.9 |
| Influenza vaccination | 70-74 | 400,143 | 71.8 |
| Influenza vaccination | 75-79 | 310,417 | 77.4 |
| Influenza vaccination | 80-84 | 237,347 | 80.3 |
| Influenza vaccination | 85-89 | 146,427 | 80.5 |
| Influenza vaccination | 90-95 | 64,238 | 79.4 |
| Influenza vaccination | 95+ | 19,826 | 76.1 |
| Pneumococcal vaccination | 65-69 | 171,814 | 38.3 |
| Pneumococcal vaccination | 70-74 | 329,895 | 59.2 |
| Pneumococcal vaccination | 75-79 | 287,843 | 71.7 |
| Pneumococcal vaccination | 80-84 | 229,129 | 77.5 |
| Pneumococcal vaccination | 85-89 | 142,058 | 78.1 |
| Pneumococcal vaccination | 90-95 | 62,385 | 77.1 |
| Pneumococcal vaccination | 95+ | 19,235 | 73.8 |
| PSA testing | 65-69 | 71,613 | 32.5 |
| PSA testing | 70-74 | 101,779 | 37.9 |
| PSA testing | 75-79 | 79,356 | 42.3 |
| PSA testing | 80-84 | 56,678 | 43.4 |
| PSA testing | 85-89 | 30,517 | 41.5 |
| PSA testing | 90-95 | 10,108 | 36 |
| PSA testing | 95+ | 1,833 | 27.2 |
| Bone density scans | 65-69 | 19,797 | 4.4 |
| Bone density scans | 70-74 | 29,459 | 5.3 |
| Bone density scans | 75-79 | 23,462 | 5.8 |
| Bone density scans | 80-84 | 16,393 | 5.5 |
| Bone density scans | 85-89 | 8,844 | 4.9 |
| Bone density scans | 90-95 | 2,544 | 3.1 |
| Bone density scans | 95+ | 393 | 1.5 |
| GP visits | 65-69 | 396,941 | 88.6 |
| GP visits | 70-74 | 511,875 | 91.8 |
| GP visits | 75-79 | 377,556 | 94.1 |
| GP visits | 80-84 | 281,987 | 95.4 |
| GP visits | 85-89 | 174,447 | 95.9 |
| GP visits | 90-95 | 77,495 | 95.7 |
| GP visits | 95+ | 24,522 | 94.1 |
| DNA primary care | 65-69 | 118,146 | 26.4 |
| DNA primary care | 70-74 | 151,958 | 27.3 |

| | | | |
|---|---|---|---|
| DNA primary care | 75-79 | 124,316 | 31 |
| DNA primary care | 80-84 | 102,601 | 34.7 |
| DNA primary care | 85-89 | 67,750 | 37.3 |
| DNA primary care | 90-95 | 29,068 | 35.9 |
| DNA primary care | 95+ | 8,057 | 30.9 |
| Low value procedures | 65-69 | 59,141 | 13.2 |
| Low value procedures | 70-74 | 87,714 | 15.7 |
| Low value procedures | 75-79 | 77,838 | 19.4 |
| Low value procedures | 80-84 | 66,686 | 22.6 |
| Low value procedures | 85-89 | 43,871 | 24.1 |
| Low value procedures | 90-95 | 18,545 | 22.9 |
| Low value procedures | 95+ | 5,086 | 19.5 |
| Low value prescriptions | 65-69 | 29 | <0.1 |
| Low value prescriptions | 70-74 | 58 | <0.1 |
| Low value prescriptions | 75-79 | 57 | <0.1 |
| Low value prescriptions | 80-84 | 40 | <0.1 |
| Low value prescriptions | 85-89 | 22 | <0.1 |
| Low value prescriptions | 90-95 | <5 | <0.1 |
| Low value prescriptions | 95+ | <5 | <0.1 |
| Hospital visit ACS condition | 65-69 | 23,948 | 5.3 |
| Hospital visit ACS condition | 70-74 | 36,985 | 6.6 |
| Hospital visit ACS condition | 75-79 | 36,772 | 9.2 |
| Hospital visit ACS condition | 80-84 | 37,255 | 12.6 |
| Hospital visit ACS condition | 85-89 | 30,822 | 17 |
| Hospital visit ACS condition | 90-95 | 17,519 | 21.6 |
| Hospital visit ACS condition | 95+ | 6,835 | 26.2 |
| Blood pressure measurement | 65-69 | 288,444 | 64.4 |
| Blood pressure measurement | 70-74 | 394,102 | 70.7 |
| Blood pressure measurement | 75-79 | 307,207 | 76.6 |
| Blood pressure measurement | 80-84 | 240,202 | 81.3 |
| Blood pressure measurement | 85-89 | 151,973 | 83.6 |
| Blood pressure measurement | 90-95 | 67,185 | 83 |
| Blood pressure measurement | 95+ | 20,893 | 80.2 |

Abbreviations: AAA: abdominal aortic aneurysm; ACS: ambulatory care sensitive; DNA: did not attend; GP: general practice; NHS: National Health Service; PSA: prostate-specific antigen.

*Broad code list: code lists were less specific e.g., screening markers could mention the relevant test, but without requiring “screen” in the code.

†Restrictive lookback: for markers with an upper age of eligibility (e.g., cancer screening and NHS health checks) the lookback period stopped at the age of upper eligibility.

### Supplementary Table 8. Phi coefficients

| | NHS health checks | Bone density scan | Bowel screen | Primary DNA | Blood pressure | Pneumococcal vaccine | Influenza vaccine | ACS conditions | Low-value procedures | GP visits |
|---|---|---|---|---|---|---|---|---|---|---|
| NHS health checks | 1 | | | | | | | | | |
| Bone density scan | 0.02 | 1 | | | | | | | | |
| Bowel screen | 0.21 | 0.02 | 1 | | | | | | | |
| Primary DNA | -0.06 | 0.03 | -0.05 | 1 | | | | | | |
| Blood pressure | -0.14 | 0.03 | -0.08 | 0.16 | 1 | | | | | |
| Pneumococcal vaccine | -0.06 | 0.03 | -0.12 | 0.07 | 0.18 | 1 | | | | |
| Influenza vaccine | -0.01 | 0.04 | -0.03 | 0.06 | 0.23 | 0.41 | 1 | | | |
| ACS conditions | -0.08 | 0.02 | -0.12 | 0.11 | 0.13 | 0.07 | 0.05 | 1 | | |
| Low-value procedures | -0.05 | 0.04 | -0.06 | 0.12 | 0.11 | 0.07 | 0.06 | 0.15 | 1 | |
| GP visits | 0.01 | 0.05 | -0.02 | 0.14 | 0.42 | 0.21 | 0.33 | 0.07 | 0.11 | 1 |

Abbreviations: ACS: ambulatory care sensitive; DNA: did not attend; GP: general practice; NHS: National Health Service.
Note: the correlations are calculated using phi coefficient for binary variables.

### Supplementary Table 9. Phi coefficients males

| | AAA screen | NHS health checks | PSA test | Bone density scan | Bowel screen | Primary DNA | Blood pressure | Pneumococcal vaccine | Influenza vaccine | ACS conditions | Low-value procedures | GP visits |
|---|---|---|---|---|---|---|---|---|---|---|---|---|
| AAA screen | 1 | | | | | | | | | | | |
| NHS health checks | 0.11 | 1 | | | | | | | | | | |
| PSA test | 0.00 | 0.03 | 1 | | | | | | | | | |
| Bone density scan | -0.01 | 0.00 | 0.06 | 1 | | | | | | | | |
| Bowel screen | 0.24 | 0.17 | 0.01 | -0.01 | 1 | | | | | | | |
| Primary DNA | -0.05 | -0.05 | 0.06 | 0.03 | -0.05 | 1 | | | | | | |
| Blood pressure | -0.04 | -0.14 | 0.12 | 0.03 | -0.06 | 0.17 | 1 | | | | | |
| Pneumococcal vaccine | -0.12 | -0.07 | 0.1 | 0.03 | -0.12 | 0.07 | 0.22 | 1 | | | | |

| | | | | | | | | | | | | |
|---|---|---|---|---|---|---|---|---|---|---|---|---|
| Influenza vaccine | -0.02 | -0.02 | 0.13 | 0.03 | -0.05 | 0.07 | 0.26 | 0.42 | 1 | | | |
| ACS conditions | -0.07 | -0.07 | 0.02 | 0.03 | -0.11 | 0.11 | 0.1 | 0.07 | 0.06 | 1 | | |
| Low-value procedures | -0.04 | -0.04 | 0.07 | 0.04 | -0.06 | 0.12 | 0.12 | 0.07 | 0.07 | 0.16 | 1 | |
| GP visits | 0.01 | 0.00 | 0.17 | 0.04 | -0.02 | 0.15 | 0.45 | 0.23 | 0.36 | 0.07 | 0.11 | 1 |

Abbreviations: AAA: abdominal aortic aneurysm; ACS: ambulatory care sensitive; DNA: did not attend; GP: general practice; NHS: National Health Service; PSA: prostate-specific antigen.
Note: the correlations are calculated using phi coefficient for binary variables.

### Supplementary Table 10. Phi coefficients females

| | Breast screen | Cervical screen | NHS health checks | Bone density scan | Bowel screen | Primary DNA | Blood pressure | Pneumococcal vaccine | Influenza vaccine | ACS conditions | Low-value procedures | GP visits |
|---|---|---|---|---|---|---|---|---|---|---|---|---|
| Breast screen | 1 | | | | | | | | | | | |
| Cervical screen | 0.11 | 1 | | | | | | | | | | |
| NHS health checks | 0.09 | 0.05 | 1 | | | | | | | | | |
| Bone density scan | 0.03 | 0.01 | 0.03 | 1 | | | | | | | | |
| Bowel screen | 0.21 | 0.06 | 0.23 | 0.04 | 1 | | | | | | | |
| Primary DNA | -0.02 | -0.02 | -0.06 | 0.03 | -0.05 | 1 | | | | | | |
| Blood pressure | 0.00 | 0.02 | -0.14 | 0.03 | -0.09 | 0.16 | 1 | | | | | |
| Pneumococcal vaccine | 0.01 | 0.06 | -0.06 | 0.03 | -0.11 | 0.07 | 0.17 | 1 | | | | |
| Influenza vaccine | 0.05 | 0.06 | 0.00 | 0.04 | -0.02 | 0.05 | 0.21 | 0.39 | 1 | | | |
| ACS conditions | -0.05 | -0.03 | -0.08 | 0.01 | -0.12 | 0.11 | 0.12 | 0.06 | 0.04 | 1 | | |
| Low-value procedures | -0.01 | 0.00 | -0.05 | 0.05 | -0.05 | 0.11 | 0.11 | 0.07 | 0.05 | 0.15 | 1 | |
| GP visits | 0.03 | 0.05 | 0.01 | 0.05 | -0.02 | 0.14 | 0.41 | 0.19 | 0.31 | 0.07 | 0.10 | 1 |

Abbreviations: ACS: ambulatory care sensitive; DNA: did not attend; GP: general practice; NHS: National Health Service.
Note: The correlations are calculated using phi coefficient for binary variables.

### Figures

**Supplementary Figure 1. Demonstration of lookback periods for markers with upper age of eligibility.** For markers with an upper age of eligibility (breast, cervical, bowel cancer screening and NHS health checks), if an individual was aged under the upper age limit at index, then we used a consistent lookback period (e.g., three years). For individuals aged over this limit at index, the lookback period would start at a consistent age (e.g., 67 years) and would extend until their index date. This decision was taken as there was a concern that for older individuals electronic data transfer would have meant that the date of transfer was recorded rather than the date of event occurrence.

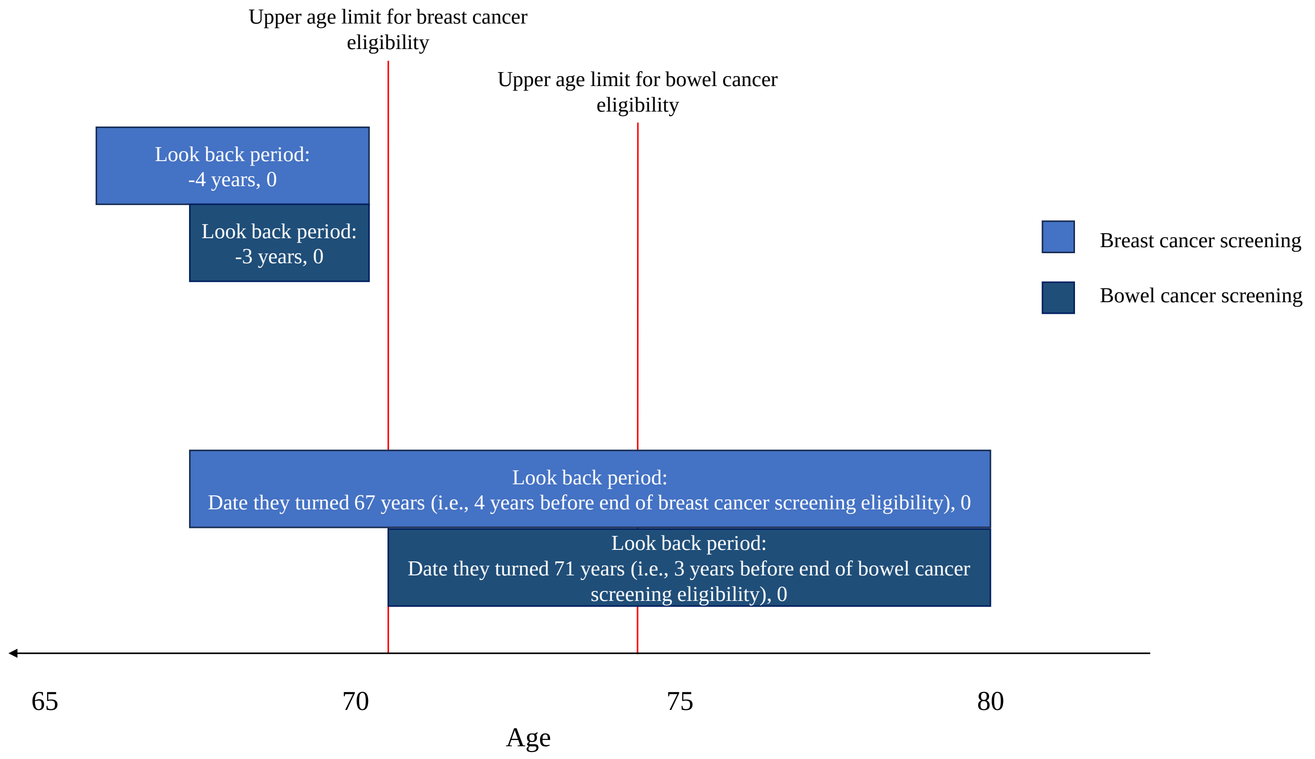


**Supplementary Figure 2. Histogram of total number of GP visits per patient in one year before index.** As majority of individuals in the study had at least one GP visit, we conducted a *post-hoc analysis* that identified the total number of GP visits per patient.

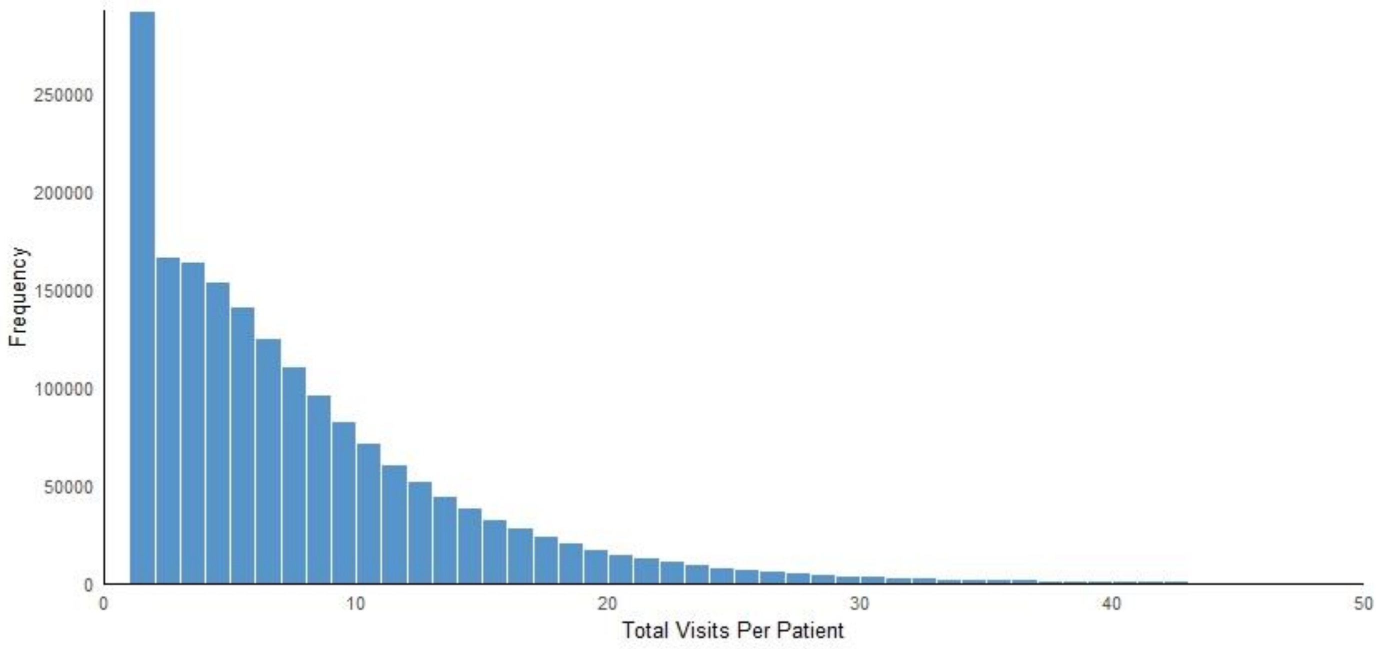


**Supplementary Figure 3: Prevalence of over the median number of GP visits, stratified by age category.** As majority of individuals in the study had at least one GP visit, we conducted a *post-hoc analysis* that identified the prevalence of over the median number of visits per year (7), stratified by age category.

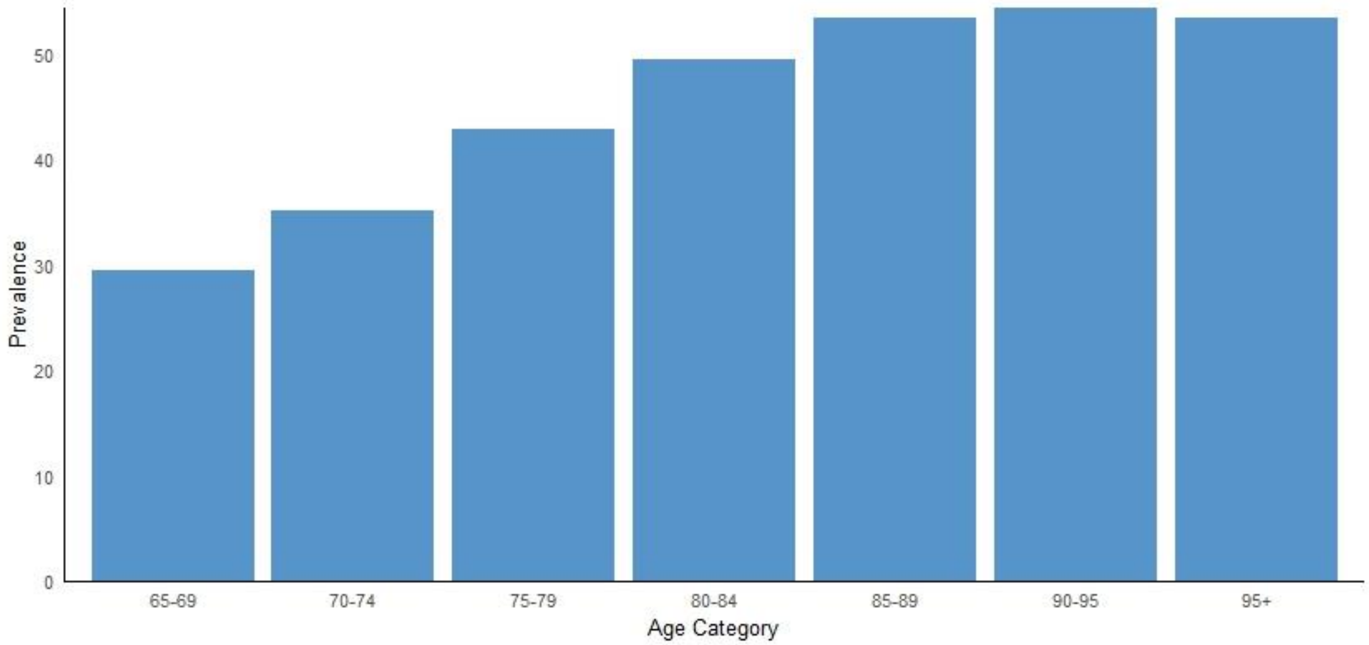
